# Transcriptomics of Type 2 Diabetic and Healthy Human Neutrophils

**DOI:** 10.1101/19011353

**Authors:** Sarah E. Kleinstein, Jamison McCorrison, Alaa Ahmed, Hatice Hasturk, Thomas E. Van Dyke, Marcelo Freire

## Abstract

**Objectives:** Chronic inflammatory diseases, including diabetes and cardiovascular disease, are heterogeneous and often co-morbid, with increasing global prevalence. Uncontrolled type 2 diabetes (T2D) can result in severe inflammatory complications. As neutrophils are essential to inflammation, we conducted RNA-seq transcriptomic analyses to investigate the association between neutrophil gene expression and T2D phenotype. Further, as specialized pro-resolving lipid mediators, including resolvin E1 (RvE1), can actively resolve inflammation, we further surveyed the impact of RvE1 on isolated neutrophils.

**Methods:** Cell isolation and RNA-seq analysis of neutrophils from N=11 T2D and N=7 healthy individuals with available clinical data was conducted. Additionally, cultured neutrophils (N=3 T2D, N=3 healthy) were perturbed with increasing RvE1 doses (0nM, 1nM, 10nM, or 100nM) prior to RNA-seq. Data was evaluated through a bioinformatics pipeline including pathway analysis and *post hoc* false-discovery rate (FDR)-correction.

**Results:** We observed significant differential expression of 50 genes between T2D and healthy neutrophils (p<0.05), including decreased T2D gene expression in inflammatory- and lipid-related genes *SLC9A4, NECTIN2* and *PLPP3* (p<0.003). RvE1 treatment induced dose-dependent differential gene expression (uncorrected p<0.05) across groups, including 59 healthy and 216 T2D neutrophil genes. Comparing T2D to healthy neutrophils, 1097 genes were differentially expressed across RvE1 doses, including two significant genes, *LILRB5* and *AKR1C1*, involved in inflammation (p<0.05).

**Conclusions:** Inflammatory- and lipid-related genes were differentially expressed between T2D and healthy neutrophils, and RvE1 dose-dependently modified gene expression in both groups. Unraveling the mechanisms regulating abnormalities in diabetic neutrophil responses could lead to better diagnostics and therapeutics targeting inflammation and inflammation resolution.

## Introduction

The increasing global prevalence of chronic inflammatory diseases, such as diabetes, is of critical concern to human health. The global prevalence of diagnosed adult diabetes was ∼415 million in 2015, and is estimated to rise to ∼642 million by 2040 (1). Type 2 diabetes (T2D) is modulated by defective metabolic and immune responses and can lead to inflammatory complications, including kidney, nerve, cardiovascular, eye, and periodontal diseases (2), with up to 60% of diabetic individuals having moderate to severe periodontitis (3). Nevertheless, the endogenous pathways that trigger and sustain unresolved low-grade inflammation in T2D are not completely understood. While acute inflammation is a protective immune response, chronic inflammatory diseases result from uncontrolled inflammation and failure of immune cells to restore homeostasis. In particular, the critical role of neutrophils in both the initiation and resolution of inflammation has become apparent, with neutrophil abnormalities a common link across chronic inflammatory diseases, including diabetes (4–8). Such neutrophil abnormalities in chronic, unresolved inflammation include impaired neutrophil adhesion, migration, chemotaxis, cytokine signaling, and phagocytosis, as well as increased neutrophil infiltration, oxidative stress and degranulation (4, 5, 9). Appropriate resolution of inflammation programs aim to restore tissue homeostasis following acute inflammation and neutrophil migration (5, 10). This active process involves key endogenous lipid ligand mediators, such as the specialized pro-resolving mediator resolvin E1 (RvE1), which can bind and transduce agonist signals through innate immune receptors on neutrophils to resolve inflammation (4, 11).

Despite considerable exploration in the genetics of T2D, primarily through genome-wide association studies (GWAS), and the identification of over 200 implicated genomic regions, to date there has been limited ability to translate genetic information into clinically actionable subtypes of this highly heterogeneous condition (12, 13). Further, identified genomic variants tend to have very small effect sizes and these genetic studies have been heavily biased by inclusion of primarily European populations. While genetics unequivocally plays an important role in T2D, with heritability estimates ranging from a low of 25% to a large twin cohort that estimated 72% heritability (14), the role of gene expression has been less studied, particularly in the context of disease-relevant cell types. Indeed, as T2D is associated with both increased adipose tissue and circulating inflammatory mediators, which induce insulin resistance and develop a state of chronic and unresolved inflammation (15), a global survey of transcriptomics provides an unbiased method to investigate functional mechanisms of T2D. Non-targeted, unbiased transcriptomics thus has the potential to elucidate novel mechanisms important for disease biomarkers and drug efficacy, especially by revealing unknown pathways of myeloid immune cells and providing dynamic molecular information important to understanding neutrophil heterogeneity in disease.

As neutrophils are essential to both initiation and resolution of inflammation and their gene expression in diabetes has not been previously studied, we conducted RNA sequencing (RNA-seq) transcriptomic analyses to investigate gene expression in human neutrophils from T2D and healthy subjects. In this study, we found that T2D individuals had higher blood glucose and more advanced periodontal disease compared to healthy individuals, consistent with worsening overall systemic health. When we investigated global neutrophil gene expression, we observed significant downregulation of several inflammatory- or lipid metabolism-related genes in T2D neutrophils. We further conducted a perturbation study to investigate the impact of exogenous RvE1 treatment on T2D and healthy neutrophil gene expression and the inflammatory cytokine signaling response. Our results demonstrate distinct trends in gene expression and cytokine level differences between T2D and healthy neutrophils that was modified by RvE1 treatment. To our knowledge, this study represents the first investigation into global neutrophil transcriptomics in type 2 diabetes.

## Materials and Methods

### Study Subjects

Subject recruitment has been described previously (11). Briefly, T2D and healthy subjects were recruited from the patient cohorts of the Center for Clinical and Translational Research at the Forsyth Institute under Forsyth Institute Institutional Review Board-approved protocols (Protocol #11-03 and #13-07). All subjects gave signed informed consent prior to study evaluations. Clinical periodontal data and peripheral venous blood were collected. The diagnosis of T2D was made by the subject’s primary care physician following American Association of Diabetes guidelines (16). Information was collected on subject demographics (age, gender, self-reported ethnicity, and self-reported smoking status), body-mass index (BMI; kg/m^2^), blood total cholesterol, blood glucose (point-of-care), percent hemoglobin A1C (HbA1c), and periodontal condition (17). HbA1c was used to determine the level of glycemic control for diabetic subjects. Neutrophil and monocyte cell counts were determined by lab assay (described below). Individuals were excluded if they were taking insulin sensitizers, nonsteroidal anti-inflammatory drugs, or antimicrobials within 3 months of study initiation. For this transcriptomics study, a total of 13 T2D and 8 healthy subjects were included for analysis, all of whom were unrelated and over 18 years of age (range: 29-70 years of age). This included 11 T2D and 7 healthy subjects in the main transcriptomics analysis of serum neutrophils, as well as 3 T2D and 3 healthy subjects whose neutrophils were cultured for perturbation experiments (1 T2D and 2 healthy subjects were included in both the main and perturbation analyses). All cell culture and transcriptomics analysis were performed at the Forsyth Institute. Data analysis and cytokine experiments were completed at the J. Craig Venter Institute.

### Human Neutrophil Isolation and Cell Culture

Human neutrophils were isolated from whole blood by Ficoll-Histopaque density-gradient centrifugation (Histopaque-1077 and Histopaque-1119; Sigma-Aldrich), as has been described previously (11). Briefly, neutrophils were isolated after isotonic lysis of RBCs and counted with a hemocytometer; neutrophil viability (>95%) was assessed using trypan blue live/dead staining (18). Monocyte exclusion was completed by plating. A separate monocyte fraction was isolated for monocyte counting. For culture experiments, isolated neutrophils were incubated with RPMI 1640 medium (Sigma-Aldrich) supplemented with 10% FBS (v/v) (Life Technologies) at 37°C. Wright-Giemsa staining was used to identify individual cell types and confirm neutrophil isolation and purity (routinely >98%) (11).

### Resolvin Perturbation Experiments

After cell isolation, cultured neutrophils from T2D subjects (N=3) and healthy controls (N=3) were treated with one of four doses of exogenous RvE1 (0nM, 1nM, 10nM, or 100nM) in RPMI and cultured for 1 hour at 37°C. Following incubation, cells were spun down and divided in half for protein and RNA-seq analyses, respectively. Cell culture supernatant was frozen at - 80°C and extracted for subsequent cytokine analyses. For the perturbation experiments, gene expression was compared between exogenous RvE1 dose-response treatmentsin 1) T2D neutrophils only, 2) healthy neutrophils only, and 3) T2D versus healthy neutrophils.

### Cytokine Analyses

Frozen (-80°C) cell culture supernatant from the perturbation experiments (N=3 T2D and N=3 healthy subject cultured neutrophils perturbed at 0nM, 1nM, 10nM, or 100nM RvE1) was brought up to room temperature and assayed using the Invitrogen human inflammation 20-plex ProcartaPlex cytokine panel (Thermo Fisher Scientific) on a Luminex 200 instrument (Luminex) in RPMI. The 20 assayed cytokines were: MIP-1α, IL-1β, IL-4, IP-10, IL-6, IL-8, IL-10, IL-12p70, IL-13, IL-17A, IFN-γ, GM-CSF, TNF-α, MIP-1β, IFN-α, MCP-1, P-Selectin, IL-1α, sICAM-1, and E-Selectin. Following manufacturer protocols, all samples were run on a plate with 7 standards (diluted 1:4) and a control (RPMI only), with all samples, standards, and controls run in duplicate (19).

Quality control (QC) steps were conducted according to manufacturer recommendations. Briefly, any standards with <70 or >130 % recovery of beads were invalidated. Samples were also checked to ensure they had a bead count of >30 beads recovered (all samples had >100 beads recovered and none were excluded at this step). Results were reported as average pg/mL for all measured cytokines following QC. Values at or below the lower limit of quantification (LLOQ) for each cytokine (based on the standard curve after QC) were reported at the LLOQ (the average value of the lowest validated standards). LLOQ for the cytokines (in pg/mL) were as follows: MIP-1α=7.18, IL-1β=8.84, IL-4=155.02, IP-10= 4.67, IL-6=29.64, IL-8=2.47, IL-10=8.37, IL-12p70=12.23, IL-13=5.61, IL-17A=30.68, IFN-γ=12.28, GM-CSF=231.09, TNF- α=9.74, MIP-1β=30.76, IFN-α=2.10, MCP-1=15.01, P-Selectin=1089.78, IL-1α=2.92, sICAM-1=445.56, and E-Selectin=2894.92.

To compare cytokine levels between groups, unpaired t-tests were used with a significance threshold of p<0.05 and no assumption of consistent standard deviation.

### RNA Sequencing Analysis

RNA-seq was conducted identically for both the serum and cell culture neutrophil experiments. Briefly, RNA was trizol extracted and quantified by Bioanalyzer (Agilent) to confirm RNA-quality (RIN score cutoff of 7.5). Library preparation was conducted using a TruSeq stranded mRNA kit (Illumina), with quality confirmed by Bioanlyzer and sequencing conducted with a Nextseq 500 high output v2 kit on a MiSeq instrument (Illumina). Resulting RNA-seq data was evaluated through a standard bioinformatics pipeline. Briefly, paired-end Illumina sequencing reads were mapped to GRCh38, after quality filtering and trimming, and quantified using STAR v.2.2.0.1 (20) with all default parameters (except “outFilterMismatchNoverLmax” set at 0.05). Reads mapped to each gene in the genome were quantified using htseq-count v.0.9.1 (21) with the following parameters: “-f bam -r pos -s yes”.

Differential gene expression analysis was conducted using edgeR v.3.18.1 (22). Genes were excluded from subsequent analysis if they did not have a count per million (CPM)>4 in at least 4 samples. The weighted trimmed mean of M-values (TMM) method (22) was used for normalization. Common dispersion was estimated with parameters method=“deviance”, robust=TRUE. Trended dispersion and tag-wise dispersion was estimated with default parameters. Gene counts were fitted with a negative binomial generalized log-linear model. Likelihood ratio tests were conducted for the specified contrasts of the coefficients, and multiple testing was subjected to a false discovery rate (FDR) correction.

Genes of interest were highlighted for visualization using R library “beanplot” (23). Heatmaps highlighting significant genes used relative abundances built using R library “vegan” (24) and rendered using R library “heatmap.2”.

### Pathway Analyses

We also conducted pathway analyses using iPathwayGuide (Advaita Bio). For the main analysis, we subjected the list of all FDR-corrected p-values to association with pathway networks using a threshold of 0.05 for statistical significance (p-value) and a log fold change (FC) of expression with an absolute value of at least 0.6. Pathways were analyzed from the Kyoto Encyclopedia of Genes and Genomes (KEGG) database (Release 84.0+/10-26, Oct 17) (25, 26) and Gene Ontology (GO) Consortium database (2017-Nov6) (27, 28), as well as investigating the network of regulatory relations from BioGRID: Biological General Repository for Interaction Datasets v3.4.154. October 25th, 2017 (29), and diseases from the KEGG database (25, 26).

Over-represented (uncorrected p<0.05) and statistically significant associations (Bonferroni or FDR-adjusted p<0.05) with pathway classifications for “disease pathways”, “biological processes”, “molecular functions”, and “cellular components” were captured. The underlying differentially expressed genes from the strongest pathway classifications were rendered and expression levels presented in heatmaps using the R library “heatmap.plus”.

For the heatmaps, additional KEGG terms of interest were selected based on previous literature regarding dental outcomes and the development of T2D (3, 8, 12). These additional terms were highlighted as part of significant networks from the iPathwayGuide analysis, such as the JAK-Stat signaling pathway. Relative abundances for the subset of genes assigned to these terms in the KEGG database (25, 26) were used for rendering to labelled heatmaps.

## Results

### Clinical Characteristics of Study Subjects

Our study population (including both the main transcriptomics and cell culture perturbation analyses) consisted of 13 T2D cases and 8 healthy controls with clinical and demographic information on: age, gender, self-reported ethnicity, self-reported smoking status, BMI, blood total cholesterol, blood glucose, percent HbA1c, periodontal condition, neutrophil cell counts, and monocyte cell counts (**Table 1**). The T2D individuals in the cohort had higher blood glucose and more advanced periodontal disease compared to healthy individuals. Diabetes diagnosis was confirmed by elevated blood glucose and HbA1c levels, with blood glucose>126mg/dl and HbA1c>6.5% indicating diabetes, consistent with clinical criteria and the literature (16); HbA1c was not measured for healthy control individuals in this cohort. T2D individuals tended to have higher cholesterol, BMI, neutrophil counts, and monocyte counts, but these factors did not reach statistical significance, while T2D individuals were significantly older than healthy individuals (p=0.007).

**Table 1.** Clinical demographics of study subjects.

|  |  | Healthy<br>(N=8) | Type 2 Diabetic<br>(N=13) | Total Cohort<br>(N=21<sup>1</sup>) | *p*-<br>value |
| --- | --- | --- | --- | --- | --- |
| **Age (mean in years ± SD)** |  | 39 (±9.63) | 58 (±11.05) | 51 (±14.04) | *0.007* |
| **Gender (no., %)** | Male | 4 (50%) | 7 (53.85%) | 11 (52.38%) |  |
|  | Female | 4 (50%) | 6 (46.15%) | 10 (47.62%) | - |
| **Ethnicity** | Caucasian | 5 (62.50%) | 4 (30.77%) | 9 (42.86%) |  |
|  | Hispanic | 0 | 1 (7.69%) | 1 (4.76%) |  |
|  | African-American | 3 (37.50%) | 8 (61.54%) | 11 (52.38%) | - |
| **Smoking Status (no., %)** | Smokers | 1 (12.50%) | 1 (7.69%) | 2 (9.52%) |  |
|  | Former Smokers | 0 | 2 (15.38%) | 2 (9.52%) |  |
|  | Never Smokers | 7 (87.50%) | 10 (76.92%) | 17 (80.95%) | - |
| **BMI (kg/m<sup>2</sup>; mean ± SD)** |  | 28.56 (±4.47) | 30.46 (±6.27) | 29.74 (±5.61) | 0.47 |
| **Blood Cholesterol (mg/dl; mean ± SD)** |  | 144.25<br>(±20.61) | 183.77 (±66.97) | 168.71<br>(±56.80) | 0.12 |
| **Blood Glucose (mg/dl; mean ± SD)** | Normal (<100mg/dl) | 96.50 (±3.54) | - | 96.50 (±3.54) |  |
|  | Pre-diabetes (100-125mg/dl) | 107 (±3.81) | - | 107 (±3.81) |  |
|  | Diabetes (>126mg/dl) | 209 | 267.77<br>(±123.38) | 263.57<br>(±119.58) | *0.003* |
| **HbA1c % (mean ± SD)** | Normal (<5.7% HbA1c) | - | - | - |  |
|  | Pre-diabetes (5.7-6.4% HbA1c) | - | 6.15 (±0.17) | 6.15 (±0.17) |  |
|  | Diabetes (>6.5% HbA1c) | - | 9.64 (±2.63) | 9.64 (±2.63) | - |
| **Periodontal Condition (no., %)** | Healthy | 8 (100%) | 0 | 8 (38.09%) |  |
|  | Mild | 0 | 3 (23.08%) | 3 (14.29%) |  |
|  | Moderate | 0 | 6 (46.15%) | 6 (28.57%) |  |
|  | Severe | 0 | 2 (15.38%) | 2 (9.52%) |  |
|  | Gingivitis | 0 | 2 (15.38%) | 2 (9.52%) | - |
| **Neutrophil Count (mil. of cells, mean ± SD)** |  | 104.88<br>(±57.23) | 123.69 (±65.68) | 116.52<br>(±61.83) | 0.51 |
| **Monocyte Count (mil. of cells, mean ± SD)** |  | 72.50<br>(±34.13) | 99.59 (±73.72) | 89.27<br>(±62.05) | 0.34 |

<sup>1</sup> N=2 healthy and N=1 diabetic subjects were included in both the serum and cell culture perturbation analyses. *p*-values calculated by unpaired t-tests (two-sided *p*-values; *italicized p*<0.05 significant). SD, standard deviation; BMI, body-mass index.

### Global Gene Expression Levels of Neutrophils in Health and Disease

We observed statistically significant differential expression of 50 genes (FDR-corrected p<0.05, **Figure 1A, Table 2, Figure S1**) between T2D and healthy neutrophils. Differentially expressed genes were primarily inflammatory- or lipid-associated and were almost universally downregulated in T2D individuals relative to healthy individuals, with the exception of *GTSCR1* (Gilles de la Tourette syndrome chromosome region, candidate 1), a non-coding RNA gene that was upregulated in T2D neutrophils.

**Table 2.**
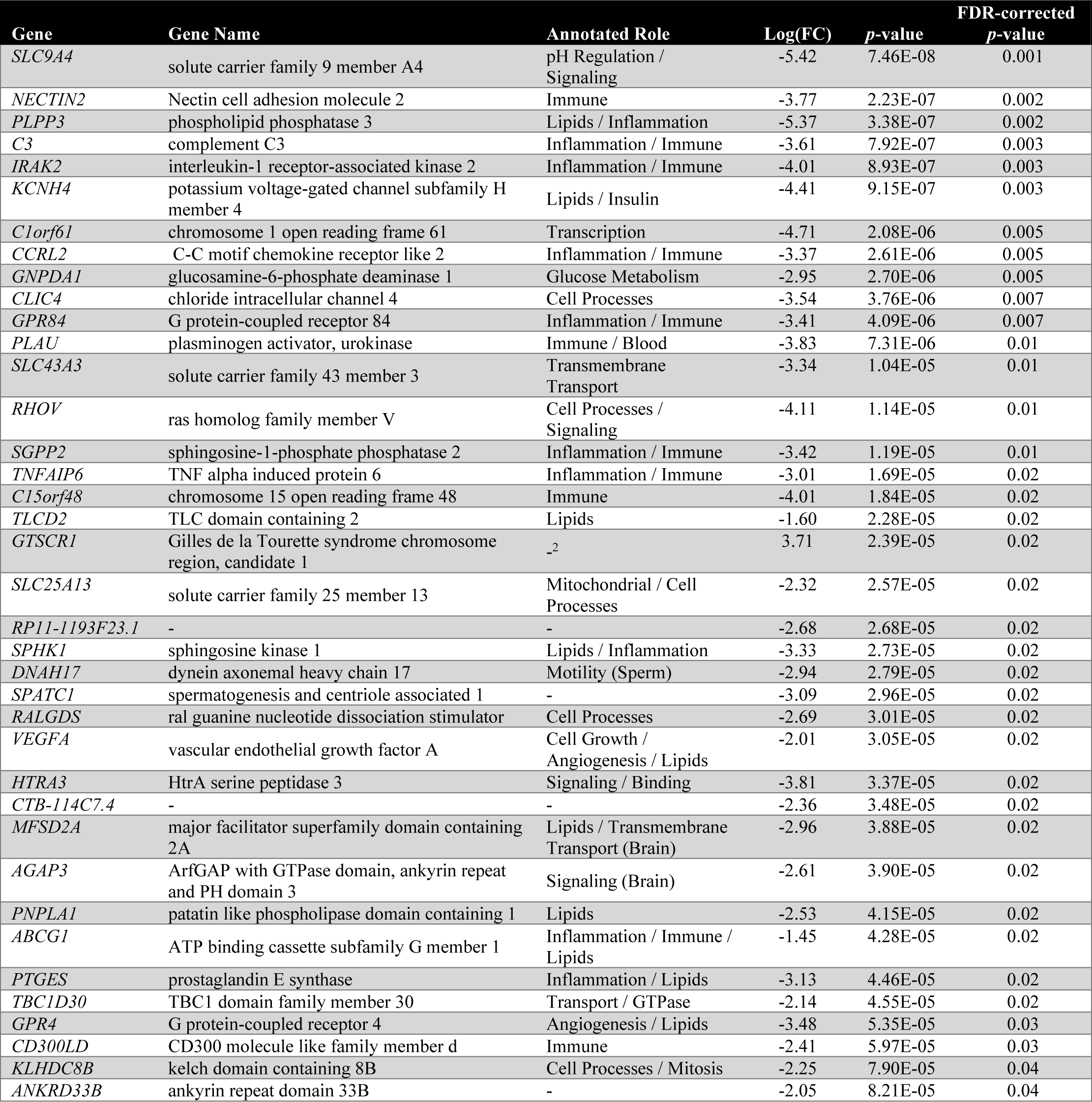
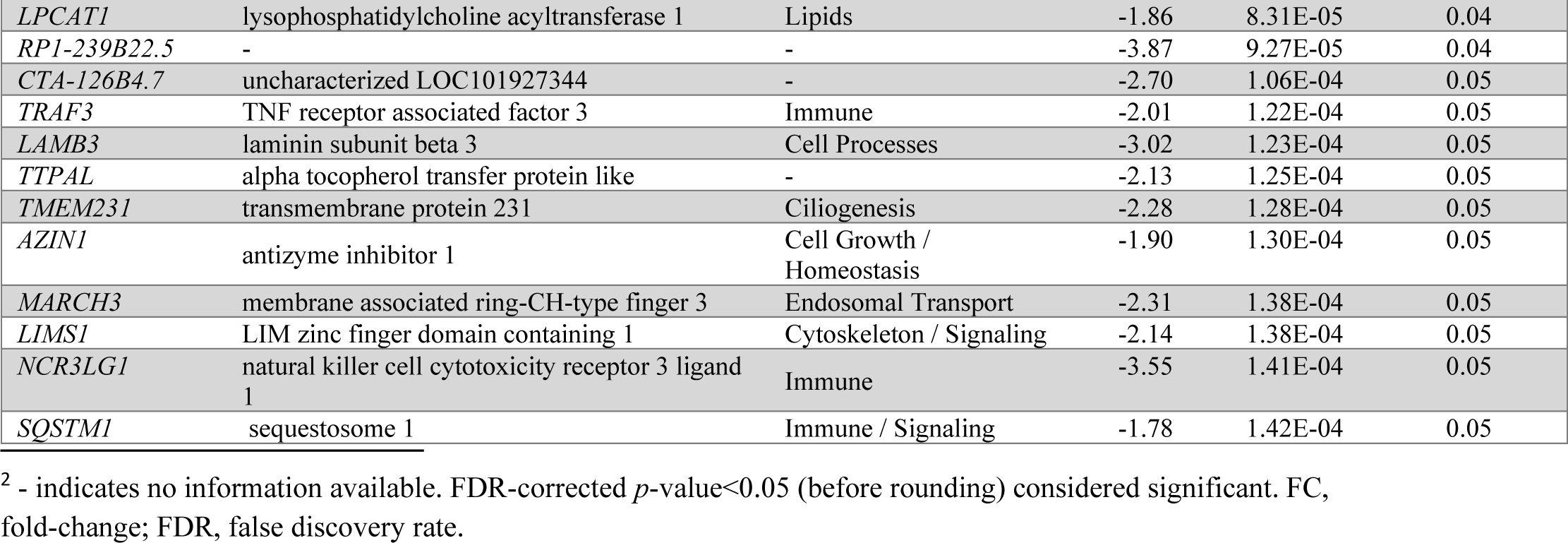
All statistically significant differentially expressed neutrophil genes comparing T2D (N=11) vs. healthy (N=7) subjects.

**Figure 1.**
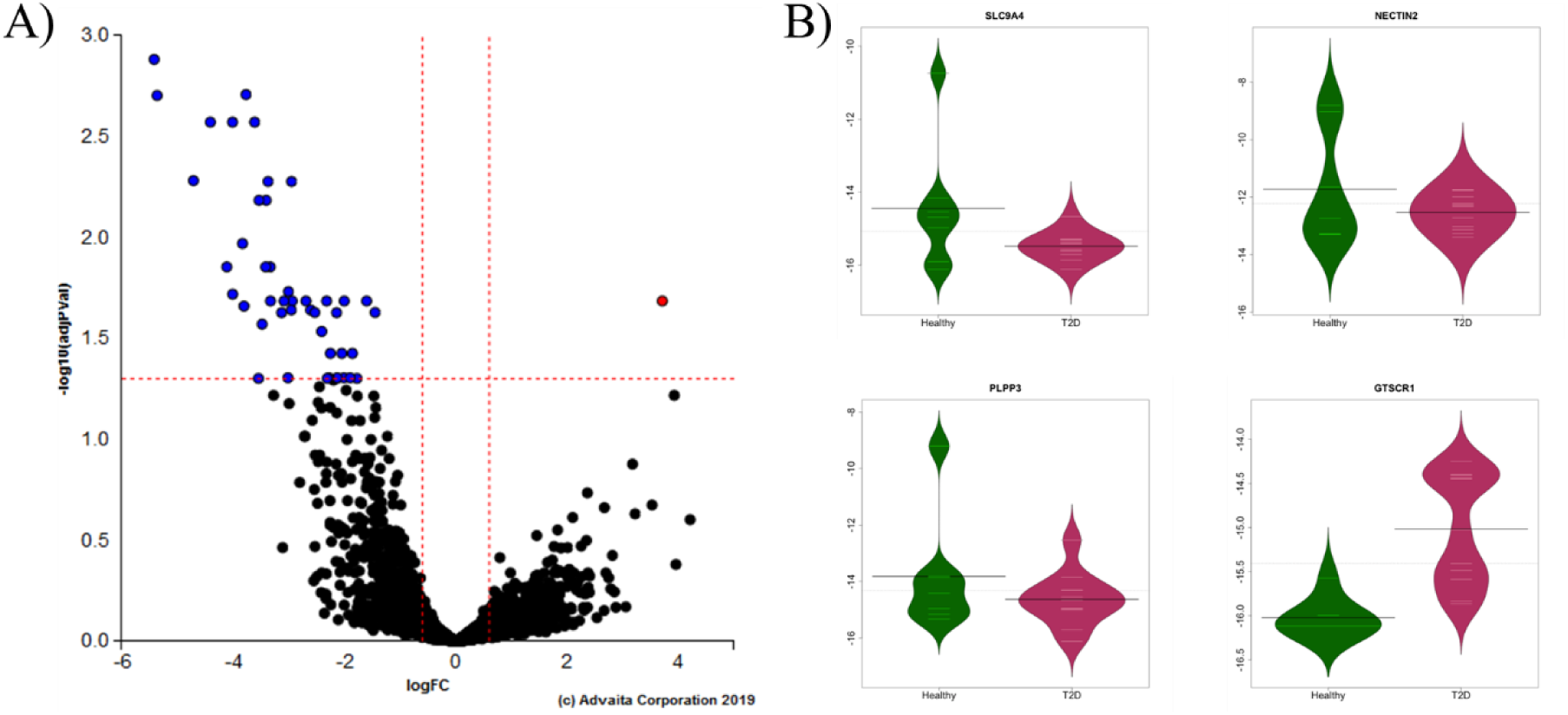
Neutrophils from type 2 diabetic subjects show down-regulated expression of inflammatory and lipid genes relative to healthy neutrophils. RNA-seq transcript expression of T2D (N=11) vs. healthy (N=7) subject neutrophils. **A)** Volcano plot of ln-transformed gene expression (x-axis) and the FDR-adjusted negative log-transformed *p*-value (y-axis). The dotted lines represent selected significant differentially expressed gene thresholds: 0.6 for fold-change, 0.05 for significance. Blue and red indicate significantly down- and up-regulated genes, respectively. **B)** Bean plots of the ln-transformed relative abundance of gene expression (line at mean) of the top 3 most significant down-regulated genes and the one significant up-regulated gene (*GTSCR1*) plotted for T2D vs. healthy individuals.

The top 3 differentially expressed genes were downregulated in T2D neutrophils (**Figure 1B**): inflammatory signaling gene *SLC9A4* (solute carrier family 9 member A4; log(FC)=-5.42, FDR-corrected p=0.001), immune regulating gene *NECTIN2* (nectin cell adhesion molecule 2; log(FC)=-3.77, FDR-corrected p=0.002), and anti-inflammatory gene *PLPP3* (phospholipid phosphatase 3; log(FC)=-5.37, FDR-corrected p=0.002); while only *GTSCR1* was upregulated in T2D neutrophils (log(FC)=3.71, FDR-corrected p=0.02, **Figure 1B**).

Of the significant differentially expressed neutrophil genes, 34% (17/50) had known immune or inflammation related roles, with 24% (12/50) of genes linked to lipid or glucose metabolism (**Table 2**). In addition to *NECTIN2* and *PLPP3*, 15 other annotated immune or inflammation associated genes had significantly lower gene expression in T2D neutrophils (**Table 2)**, while *KCNH4, GNPDA1, TLCD2, VEGFA, MFSD2A, PNPLA1, GPR4*, and *LPCAT1* showed lower expression in T2D neutrophils and had previously been linked to lipid or glucose metabolism. Indeed, several significant differentially expressed genes, including *PLPP3, SPHK1, ABCG1*, and *PTGES*, have previously been indicated to have both lipid- and inflammatory-related roles.

This interaction between lipid and inflammatory pathways was further evident when we investigated neutrophil gene expression by biological pathways and observed that most of the significant biological pathways were lipid-related, despite the abundance of immune- and inflammatory-related genes among our significant results (**Table 3, Figures S2-S3**).

**Table 3.**
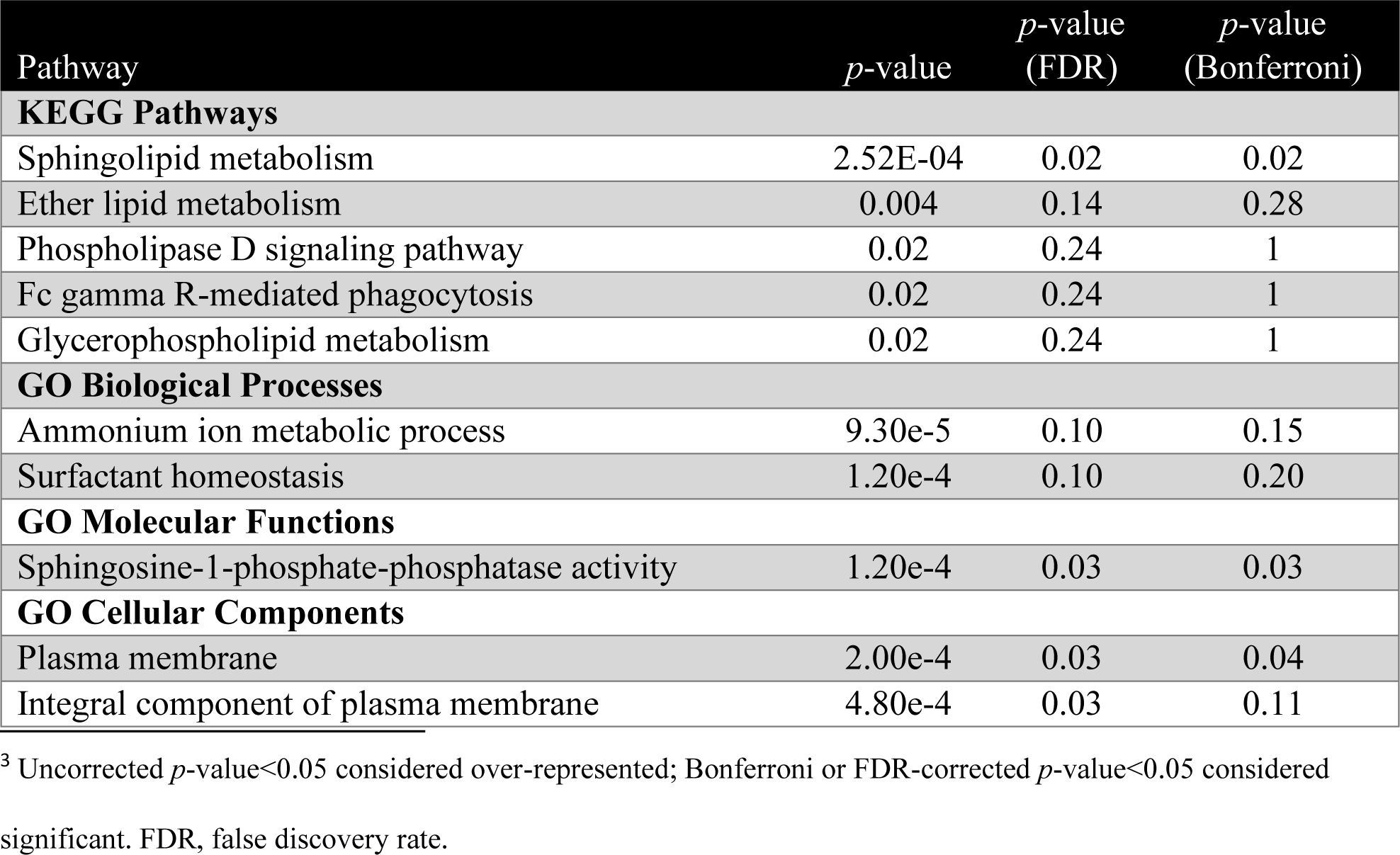
Top over-represented pathways included in pathway analyses.^3^

Overall, 13 pathways and 19 diseases were over-represented (uncorrected p<0.05) in our cohort by gene expression differences. The top 5 over-represented KEGG pathways were heavily lipid-related: sphingolipid metabolism, ether lipid metabolism, phospholipase D signaling pathway, Fc gamma R-mediated phagocytosis, and glycerophospholipid metabolism; though only sphingolipid metabolism was statistically significant after Bonferroni or FDR correction (corrected p=0.02, **Figure S3**). Of the 19 over-represented diseases, 15 remained significant by FDR correction, but none remained significant after Bonferroni correction, and all 19 had only a single differentially expressed gene represented in that particular disease pathway. The top 5 disease pathways were: citrullinemia, Paget’s disease, endometriosis, junctional epidermolysis bullosa, and avascular necrosis/osteonecrosis of femoral head. Additionally, top GO biological process, molecular function, and cellular component pathways were interrogated (27, 28). Though none reached significance after Bonferroni or FDR correction, the top GO pathways also included lipid-related and plasma membrane pathways (**Table 3**).

When we investigated the top KEGG and GO biological pathways (**Table 3**) by individual gene expression across the top 50 genes, we observed hierarchical clustering of pathway and gene expression by groups of individuals (**Figure 2**). These clusters primarily separated out diabetic and healthy individuals and included disease heterogeneity in gene levels.

**Figure 2.**
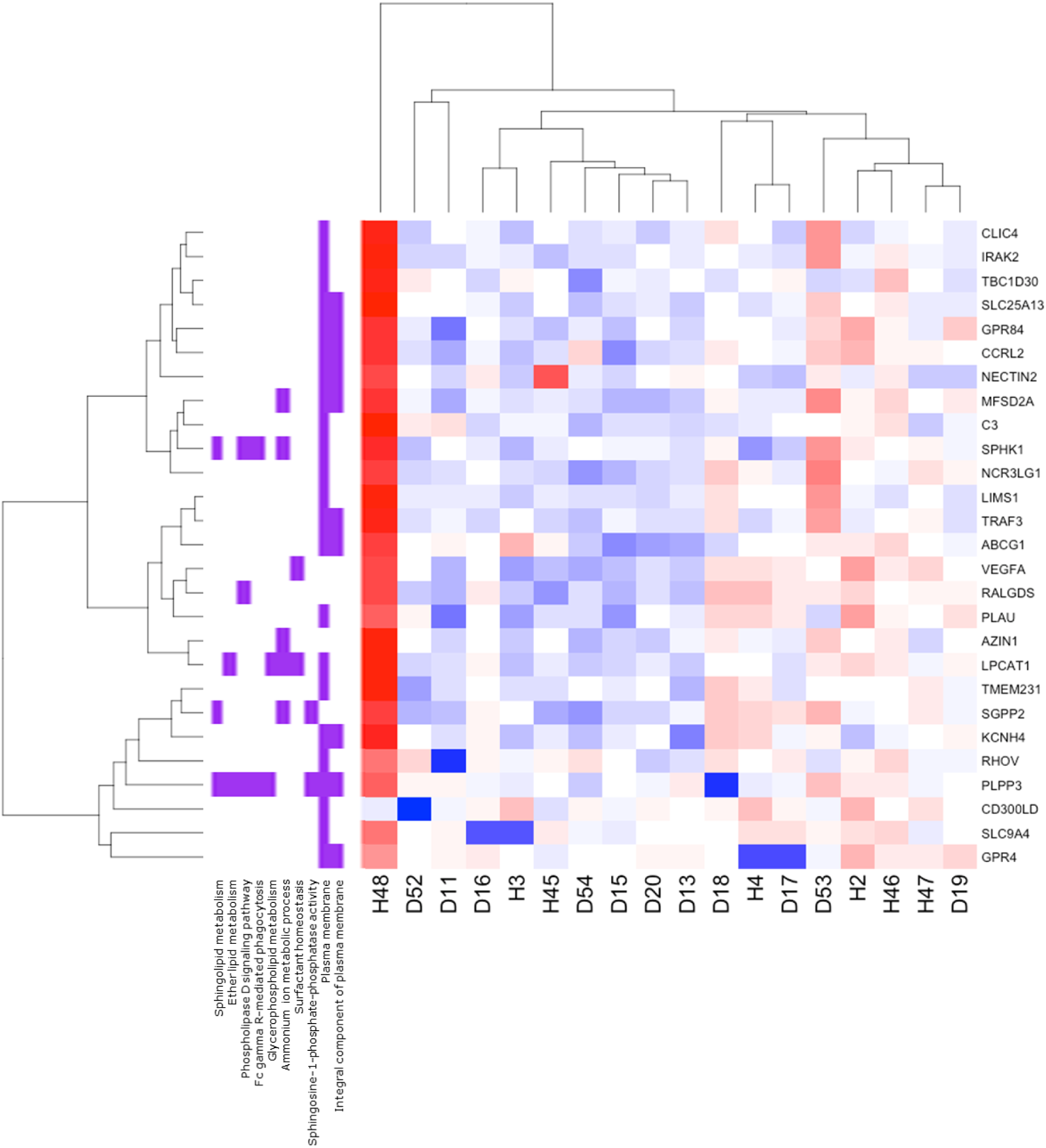
Neutrophil gene expression in top biological pathways is primarily clustered by diabetic vs. health status. RNA-seq transcript expression of T2D (D; N=11) vs. healthy (H; N=7) subject neutrophils. Heatmap of T2D and healthy gene expression by over-represented KEGG and Gene Ontology (GO) pathways and top 50 genes (FDR-corrected p<0.05) in those pathways. Blue indicates decreased and red indicates increased gene expression.

### Neutrophil Gene Expression by Lipid Ligand Resolvin E1 Dose-Response

Because endogenous inflammatory pathways are deficient in chronic inflammatory diseases like T2D, we aimed to understand whether treating isolated neutrophils with an exogenous lipid ligand resolution mediator, RvE1, would impact transcription phenotypes.

We conducted a perturbation experiment across a range of clinically relevant RvE1 doses (0-100nM) and observed that, as expected, dosing of the RvE1 ligand impacted T2D neutrophils differently than healthy neutrophils. We investigated this dose-response phenotype between three comparisons: 1) T2D neutrophils only, 2) healthy neutrophils only, and 3) T2D vs. healthy neutrophils.

RvE1 treatment induced differential gene expression (uncorrected p<0.05) across doses (**Figure 3**), including 59 genes in healthy and 216 genes in T2D neutrophils. Comparing T2D to healthy neutrophils, 1097 genes were differentially expressed across treatment doses, including two statistically significant (FDR-corrected p<0.05) inflammatory genes, *LILRB5* (leukocyte immunoglobulin like receptor B5) and *AKR1C1* (aldo-keto reductase family 1 member C1). Neutrophils from T2D subjects had a stronger response to RvE1 treatment dose-dependently (more differentially expressed genes), particularly at the clinically relevant dose, 100nM (11) (117 differentially expressed genes between100nM and 10nM RvE1), showing modified gene expression by RvE1 treatment. In contrast, healthy neutrophils were not as perturbed by RvE1 dosing (few differentially expressed genes between doses), though some effect was observed at the 10nM dose (25 differentially expressed genes between 10nM and 0nM RvE1). Treatment of T2D neutrophils with 100nM RvE1 resulted in a reduction in the number of differentially expressed genes in T2D neutrophils compared to healthy neutrophils (98 genes when T2D neutrophils were treated with 100nM RvE1 versus 169 genes without RvE1 treatment, 0nM).

**Figure 3.**
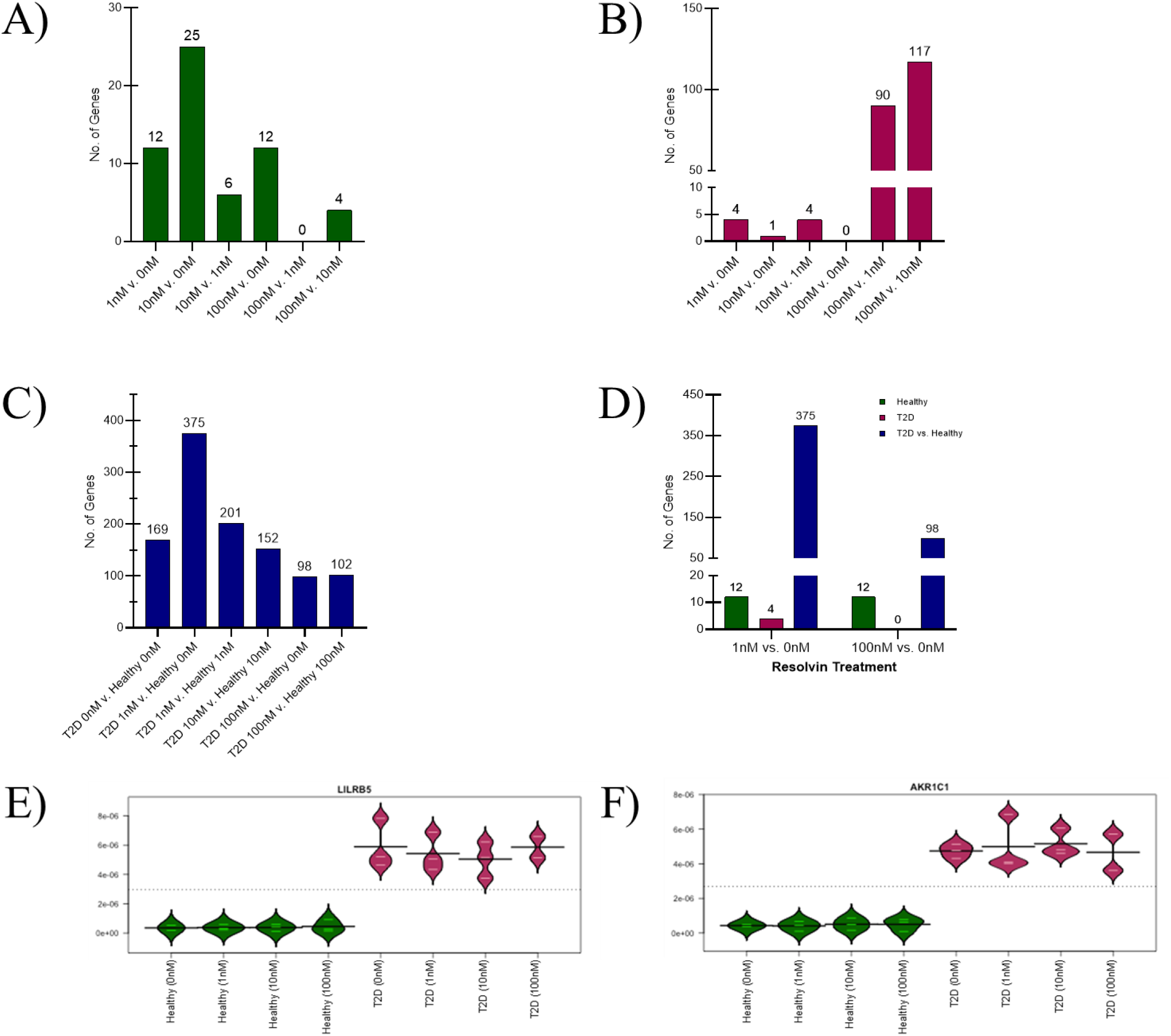
Resolvin E1 treatment has a stronger impact on gene expression in T2D relative to healthy neutrophils. Number of genes differentially expressed (uncorrected p<0.05) by resolvin E1 (RvE1) treatment dose (0nM, 1nM, 10nM, or 100nM) across groups: **A)** healthy only, **B)** T2D only, **C)** T2D (higher dose) vs. healthy (lower dose) in cultured neutrophils (N=3 T2D, N=3 healthy). **D)** Number of genes differentially expressed (uncorrected p<0.05) in healthy, T2D and T2D vs. healthy groups, comparing low (1nM vs. 0nM) to high (100nM vs. 0nM) RvE1 dose. **E-F)** Bean plots of the relative abundance of gene expression (line at mean) of the two significant differentially expressed genes (*LILRB5* and *AKR1C1*; FDR-corrected p<0.05) plotted for T2D vs. healthy individuals. T2D, type 2 diabetes.

An interesting observation was that, in general, the differentially expressed neutrophil genes across RvE1 doses tended to be unique to each comparison: 1) T2D, 2) healthy, or 3) T2D vs. healthy groups. Within each comparison the genes differentially expressed remained mostly consistent across RvE1 doses. However, the genes differentially expressed between T2D and healthy neutrophils in the cell culture perturbation model without RvE1 treatment were generally not the same as the significant differentially expressed serum neutrophil genes observed in the main analysis, with the exception of *NECTIN2, HTRA3*, and *ABCG1*, which were differentially expressed in both serum and cell culture. Interestingly, when we perturbed T2D neutrophils with 100nM RvE1 and did not perturb healthy neutrophils, only *HTRA3* remained strongly differentially expressed (p<0.05), while *NECTIN2* and *ABCG1* showed less differential expression. Overall, there were distinct trends in neutrophil responses to RvE1 perturbation between diabetic and healthy neutrophils in cell culture.

### Diabetic and Healthy Neutrophil Cytokines Respond Differently to Resolvin E1 Perturbation

To confirm that gene expression changes influenced neutrophil protein expression, we analyzed cytokine production of neutrophil cell culture supernatants following the same RvE1 dose-response studies. Results showed trends of differential inflammatory cytokine concentrations between T2D and healthy neutrophils and across RvE1 doses. Out of a panel of 20 human cytokines, the concentration of 8 cytokines (MIP-1α, IL-4, IL-8, MIP-1β, P-Selectin, sICAM-1, TNF-α, and IL-1α) were within the range of quantification and were included in the analysis (**Figure 4, Table S1**). Without any RvE1 perturbation there were differences in levels of these cytokines between T2D and healthy cell cultured neutrophils, with T2D neutrophils having higher TNF-α and P-Selectin levels but lower MIP-1β, IL-8, and sICAM-1 levels (**Figure 4A**). Across all RvE1 doses, P-Selectin and sICAM-1 levels remained consistently higher, while IL-8 levels remained lower in T2D compared to healthy neutrophils. However, at 1nM RvE1 treatment, MIP-1β levels in T2D neutrophils rose to the baseline levels of healthy neutrophils, while healthy neutrophil levels stayed consistent. At higher RvE1 doses, including 10nM and 100nM, MIP-1β levels rose in both T2D and healthy neutrophils; there was a stronger, dose-dependent increase among healthy neutrophils. After treatment with 10nM or 100nM RvE1, TNF-α levels were elevated in healthy neutrophils, reaching levels seen in T2D neutrophils; a similar effect was observed at 100nM RvE1 for P-Selectin, though healthy levels (1511pg/mL) did not rise to fully match T2D levels (1751pg/mL). We also observed some distinct neutrophil cytokine profiles in individual subjects, consistent with known inter-individual cytokine variation. The cytokine level differences did not reach statistical significance at this sample size. Overall, we observed trends of several potentially interesting differences in cytokines between T2D and healthy neutrophils, indicating that both diabetic and healthy neutrophils may have a distinct functional signaling response to RvE1 treatment in a dose-dependent manner.

**Figure 4.**
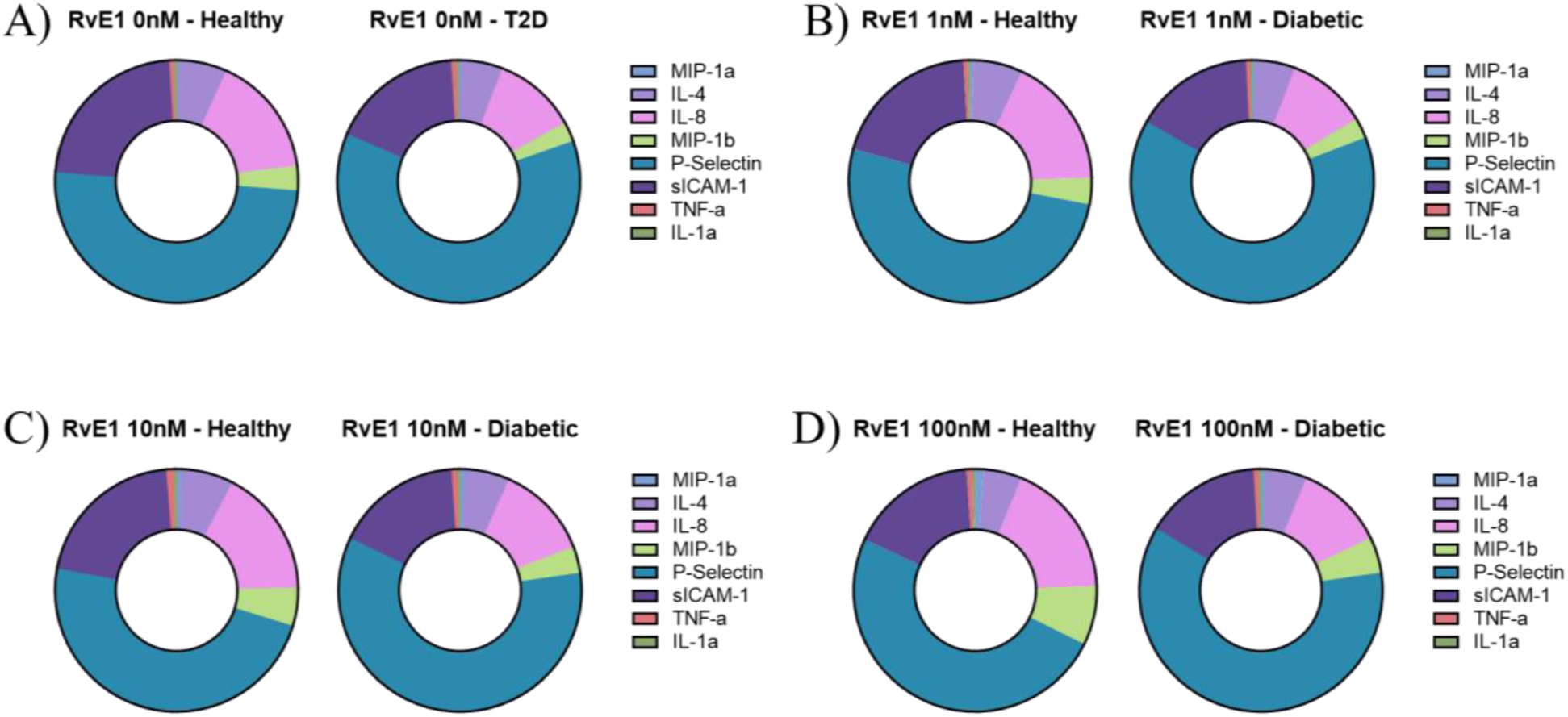
Type 2 diabetic and healthy neutrophils showed differential inflammatory cytokine concentrations across resolvin E1 doses. Human inflammatory cytokine levels (cytokines: MIP-1α, IL-4, IL-8, MIP-1β, P-Selectin, sICAM-1, TNF-α, IL-1α; average pg/mL) in response to resolvin E1 (RvE1) dose, comparing T2D (N=3) to healthy (N=3) cultured neutrophils with: **A)** vehicle only (0nM), **B)** 1nM, **C)** 10nM, or **D)** 100nM RvE1 treatment. For each cytokine, values at or below the lower limit of quantification (LLOQ; based on a 7-standard curve at 1:4 dilution) were reported as the LLOQ value.

## Discussion

We conducted the first study of neutrophil gene expression in the chronic inflammatory disease type 2 diabetes. We observed differential expression between T2D and healthy neutrophils of several inflammatory- or lipid-associated genes, indicating the potential for distinct transcriptomic profiles between these groups among a key and novel immune cell type. Further, as many of the significant differentially expressed genes had direct lipid- or glucogenesis-related roles, neutrophil transcriptomics may provide a window directly into relevant disease pathogenesis beyond their role in immune or inflammatory genes.

The top genes significantly down-regulated in T2D neutrophils compared to healthy neutrophils tended to be biologically relevant across inflammatory- and lipid-associated gene pathways and included *SLC9A4, NECTIN2*, and *PLPP3*.

*SLC9A4* is a plasma membrane solute carrier protein that acts in homeostatic pH regulation and proton transport, including to eliminate metabolism-generated acids and absorb sodium, with an important role in signal transduction (30, 31). *SLC9A4* is in a genomic region with genes involved in the IL-1 receptor (IL-1R) and IL-18 pathways (32, 33). This *SLC9A4/IL-1R/IL-18* locus has important roles in cytokine signaling and inflammatory response (32, 33). The IL-18 pathway induces synthesis of the cytokine IFN-γ in T cells, which has been shown to act in mucosal inflammation in celiac disease (32, 34). Celiac disease is a chronic inflammatory and autoimmune disease that shares several genetic risk loci with type 1 diabetes (32). A celiac disease GWAS identified associations with the *SLC9A4/IL-1R/IL-18* locus, and a variant in this locus further showed an allelic dosage effect that reduced *IL18RAP* mRNA expression in celiac patients, indicating a direct link between this locus’ genotype and biologically relevant gene expression in a chronic inflammatory disease (32). This reduced IL-18 cytokine expression in celiac disease is consistent with the overall reduced expression of *SLC9A4* observed among diabetic neutrophils in our study. Another GWAS linked *SLC9A4* with regulation of IL-33/ST2 (suppression of tumorigenicity 2, part of the IL-1R family) signaling, which has previously been implicated in both immune and inflammatory diseases, including cardiovascular disease (33). In addition, a variant in *SLC9A4* was associated with eosinophil, though not neutrophil, cell counts in a GWAS of white blood cell subtypes utilizing subjects from the BioBank Japan Project, further indicating a direct role for *SLC9A4* in immune cell activity (35).

*NECTIN2* is a cholesterol responsive, adherens junction plasma membrane glycoprotein (36, 37). *NECTIN2* is also involved in extravasation and angiogenesis, and may act at vascular inflammation sites to regulate transendothelial leukocyte migration (36, 37). This study is the first exploration of *NECTIN2* expression in neutrophils. We observed downregulation of *NECTIN2* among T2D subjects, which is consistent with a continuous cell migration phenotype. Further, we observed higher cholesterol levels among our T2D subjects, consistent with both the role of *NECTIN2* and the known propensity for high cholesterol in diabetic patients. Previous evidence has shown that *NECTIN2* also regulates immune system responses, including modulating T cell signaling and viral pathogenesis (38). Further, *NECTIN2* has shown potential associations with several diseases, including chronic inflammatory diseases.

*PLPP3* is a plasma membrane adherens junction phospholipid phosphatase that converts phosphatidic acid to diacylglycerol and has roles in lipid metabolism and Wnt signaling (30, 39, 40). The Wnt signaling pathway is involved in a menagerie of cellular functions and development, and its dysfunction has been linked to diseases including cancer, metabolic syndrome, diabetes, and diabetic neuropathy (41). It has also previously been demonstrated that silencing *PLPP3* in an endothelial cell line enhanced inflammatory cytokine secretion, leukocyte adhesion, cell survival, and migration, while its overexpression reversed those effects and instead induced apoptosis (42). This is consistent with our observation of reduced *PLPP3* expression among T2D neutrophils, as *PLPP3* normally acts in a protective role as a negative regulator of inflammatory cytokines and leukocyte adhesion. In addition, GWAS have previously associated variants in *PLPP3* with several chronic inflammatory diseases, including coronary artery disease (43–46) and eosinophilic esophagitis (47). Thus, the lipid regulatory and anti-inflammatory roles of *PLPP3* in chronic inflammatory diseases lend further evidence for a metabolic-immune axis in diabetes (8).

It is worth note that our study of neutrophil transcriptomics found that most significant gene expression was downregulated in T2D neutrophils compared to healthy neutrophils. It has traditionally been thought that gene expression in diabetic individuals would be increased, particularly among inflammatory and immune genes, and indeed gene expression studies of pancreatic islet cells showed increased gene expression in diabetes (48). However, as neutrophils experience severe dysregulation in individuals with chronic inflammatory disease (4, 5, 9), we do not believe it is surprising to see a downregulation of many key immune and inflammatory genes. Diabetic neutrophils are known to exhibit impaired immune responses and cell migration (4, 5, 9), and we observed a marked reduction in neutrophil chemotaxis in T2D compared to healthy neutrophils (data not shown), consistent with the downregulated gene expression observed in this study. Further, a recent study in adipose tissue showed that apolipoprotein M expression was decreased in T2D and obese individuals compared to lean individuals (49), further indicating a role for downregulation of genes in diabetes pathogenesis.

When we further explored neutrophil gene expression by biological pathways, we observed strong links between lipid- and inflammatory-associated genes and pathways, further emphasizing the role of the metabolic-immune axis in diabetes (8). Plasma membrane cellular pathways were also over-represented, consistent with the roles of our top differentially expressed genes. Hierarchical clustering of over-represented and biologically relevant pathways by individual expression of significant differentially expressed genes in these pathways tended to sort individuals by disease status. However, disease heterogeneity could be observed in the clustering, with some T2D individuals in a healthy cluster and vice versa, indicating the potential to identify subsets of disease severity through neutrophil transcriptomics.

To investigate the functional activity of T2D and healthy neutrophils and whether diabetic neutrophil dysregulation could be restored to a healthy, rather than diseased, phenotype, we treated cell cultured neutrophils from T2D and healthy individuals with the small lipid ligand resolution mediator RvE1. We observed that RvE1 dose-dependently modified both T2D and healthy neutrophil gene expression in cell culture, suggesting a tendency for RvE1 treatment to shift neutrophils toward a healthy phenotype. However, across treatment doses, only two genes were significantly differentially expressed between T2D and healthy neutrophils: *LILRB5* and *AKR1C1*.

*LILRB5* is an immune system gene that can bind major histocompatibility complex (MHC) class I molecules on antigen-presenting cells, inhibiting stimulation of an immune response (50). *AKR1C1* catalyzes the reaction of progesterone to its inactive form but may also regulate inflammatory cytokine signaling pathways (51), consistent with some altered cytokine concentrations observed in this study. As *LILRB5* and *AKR1C1* remained significantly differentially expressed regardless of RvE1 treatment dose, they may reflect molecular changes in neutrophil inflammatory profiles.

In general, the genes differentially expressed between untreated T2D and healthy neutrophils differed between the human serum and cell culture models. This may be due to *in vivo* vs. *ex vivo* neutrophil differences, inter-individual variation in neutrophil responses, as not all individuals whose neutrophils were cultured were also included in the baseline serum neutrophil analysis, or they may represent different genes targeting similar pathways. Indeed, when we investigated the top KEGG and GO pathways associated with differential gene expression between T2D and healthy cell cultured neutrophils with a low (1nM) dose of RvE1, the top pathways were primarily immune related, though there was less of a lipid pathway role (data not shown).

Interestingly, three genes were differentially expressed in both neutrophil cell culture without RvE1 treatment and the baseline serum analysis, including *NECTIN2, HTRA3*, and *ABCG1*. Importantly, treatment of the T2D neutrophils in cell culture with 100nM of exogenous RvE1 was able to reduce the level of differential expression of *NECTIN2* and *ABCG1*, suggesting a partial normalization of these neutrophils with RvE1 treatment.

We then used cytokine profiling to further interrogate the functional effect of RvE1 treatment on T2D and healthy neutrophils. We observed trends in different in cytokine profiles between T2D and healthy neutrophils both with and without RvE1 treatment. In general, T2D neutrophils tended to have a stronger response to RvE1 treatment, particularly at higher, clinically relevant, RvE1 doses. Without any RvE1 treatment, we observed higher levels of pro-inflammatory cytokines P-Selectin and TNF-α among T2D compared to healthy neutrophils, though levels of pro-inflammatory MIP-1β, IL-8, and sICAM-1 cytokines were decreased in T2D neutrophils. As cytokines have complex and often overlapping roles in pro- and anti-inflammatory pathways, the relative contribution of each cytokine to neutrophil dysfunction in T2D will require further investigation. We also observed differences in cytokine profiles by subject, consistent with known inter-individual variation (52). Our study was limited by its small sample size, which made more detailed subgroup analysis not feasible. However, even at this sample size, this novel cell type has shown distinct differences in transcriptomic and functional cytokine profiling between T2D and healthy individuals, including following RvE1 treatment, indicating that larger follow-up studies are warranted. Importantly, our cytokine profiling experiments showed that both T2D and healthy neutrophils were viable and capable of both producing cytokines and responding to RvE1 treatment, demonstrating the importance of concentration-based therapeutics for exogenous lipid ligands.

Key strengths of our study include the novel exploration of a biologically relevant cell type (neutrophils) in chronic inflammatory diseases, investigations of both gene level transcriptomics and pathway analyses, and the further exploration of the impact of a key molecule in inflammation resolution (RvE1) on gene expression and functional cytokine levels.

Overall, we showed that neutrophils may act differently in individuals with chronic inflammatory diseases, specifically diabetes, compared to healthy individuals (**Figure 5**). Further validation of proteomic differences between diabetic and healthy individuals could elucidate important inflammation mechanisms and potential pathways related to resolvin treatment effects, including in subgroups of disease with clinically relevant diabetes comorbidities, such as those with cardiovascular disease or periodontitis. Unraveling these mechanisms of neutrophil dysregulation in chronic inflammatory diseases could ultimately elucidate inflammation and resolution targets for better diagnostic and therapeutic treatment options.

**Figure 5.**
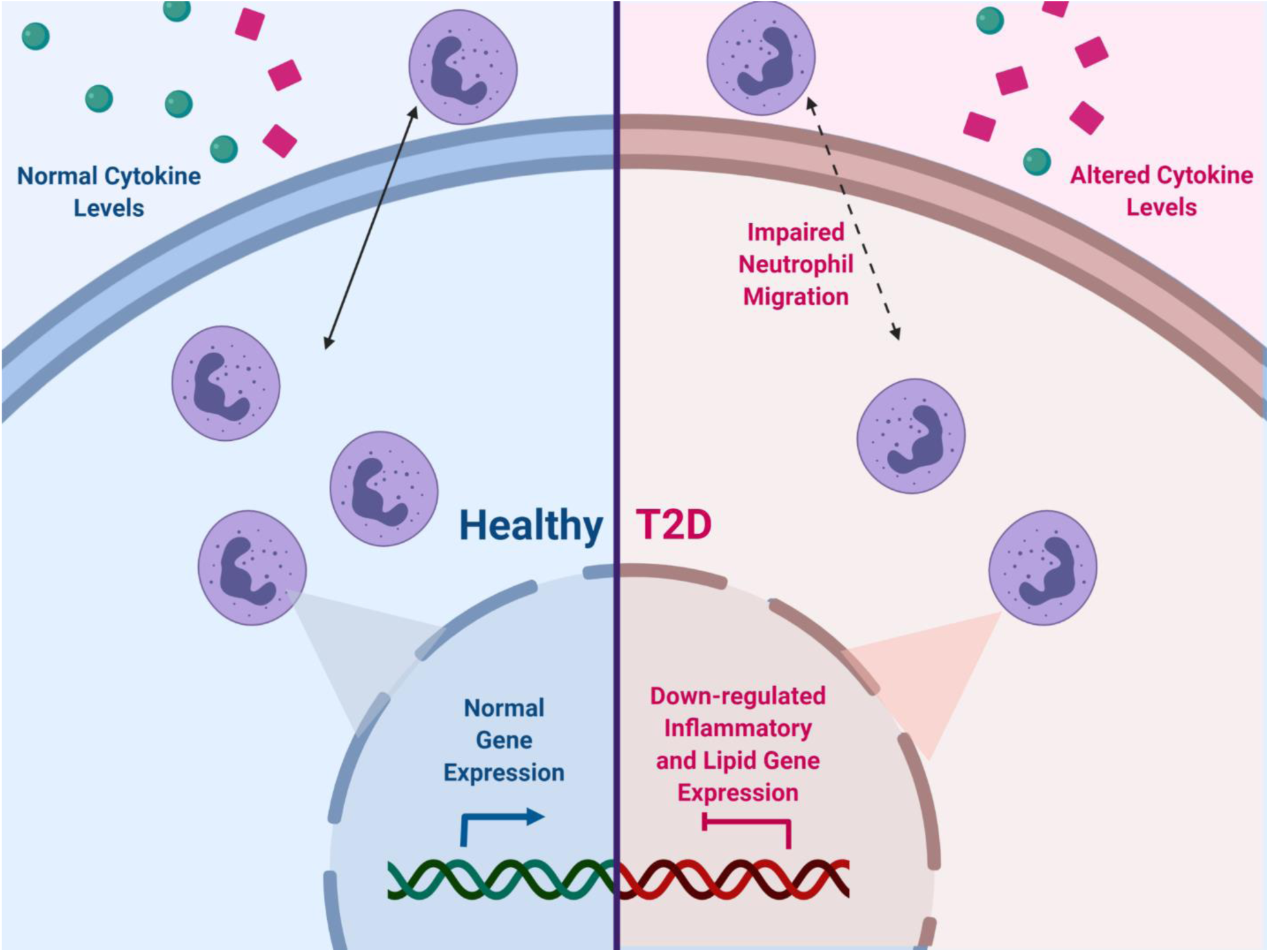
Down-regulated inflammatory- and lipid-related gene expression in type 2 diabetic neutrophils leads to overall neutrophil dysregulation, including impaired chemotaxis and altered cytokine production. Cartoon of proposed dysregulation effects in inflamed tissues in T2D (right, red) vs. healthy (left, blue) neutrophils. Created with BioRender.com. T2D, type 2 diabetes.

## Data Availability

All relevant data is available in the manuscript and supplemental material. RNA-sequencing data will be deposited in an approved database.

## Acknowledgements

We thank Tsute Chen for excellent technical assistance in transferring the transcriptomics data to JCVI for bioinformatic analyses. This work was initially performed at the Forsyth Institute and completed at the J. Craig Venter Institute.

## Abbreviations

BMI: Body-mass index
CPM: Count per million
E-selectin: Endothelial-selectin
FDR: False-discovery rate
GO: Gene Ontology
GWAS: Genome-wide association studies
HbA1c: Hemoglobin A1C
IL-1R: IL-1 receptor
IP-10: Interferon gamma-induced protein 10
KEGG: Kyoto Encyclopedia of Genes and Genomes
LLOQ: Lower limit of quantification
P-Selectin: Platelet-selectin
QC: Quality control
RNA-seq: RNA sequencing
RvE1: Resolvin E1
sICAM-1: Soluble intercellular adhesion molecule-1
ST2: Suppression of tumorigenicity 2
T2D: Type 2 diabetes
TMM: Trimmed mean of M-values

